# Prevalence of anxiety, depression, and stress among teachers during the COVID-19 pandemic: Systematic review

**DOI:** 10.1101/2021.05.01.21256442

**Authors:** David Franciole Oliveira Silva, Ricardo Ney Oliveira Cobucci, Severina Carla Vieira Cunha Lima, Fábia Barbosa de Andrade

## Abstract

**Objective:** Identifying the prevalence of anxiety, depression, and stress among teachers during the COVID-19 pandemic.

**Methods:** Systematic review of original studies published in any language. Protocol published in PROSPERO under number CRD42021240543. The search was carried out in the Web of Science, PsycINFO, Pubmed, Embase, LILACS, and SciELO databases, using the descriptors: anxiety, depression, stress, teacher, faculty, COVID-19, and their synonyms. Narrative synthesis was carried out in line with the synthesis without meta-analysis (SWiM) in systematic reviews.

**Results:** Of the 1,372 records identified, six studies, all cross-sectional, were included in the review. The studies were carried out in China, Brazil, the United States of America, India, and Spain. Five studies included more women than men. The participants were aged from 24 to 60 years. Three studies included only school teachers, two included schools and universities teachers, and one only university teachers. Of the five studies, all dealt with remote activities and only one included teachers who returned to face-to-face classes one to two weeks ago. The prevalence of anxiety ranged from 10% to 49.4%, and depression from 15.9% to 28.9%, being considerably higher in studies with teachers who worked in schools. The prevalence of stress ranged from 12.6% to 50.6%.

**Conclusions:** The prevalence of anxiety, depression, and stress was high among teachers during the pandemic, with great variation between studies. Anxiety and stress were more prevalent in the Spanish study. The results show the need for measures for the care of teachers’ mental health, especially when returning to face-to-face classes.

**What is already known about this subject?:**

➢ With remote classes during the COVID-19 pandemic, there were changes in the professional practice of teachers.
➢ Sudden changes in professional practice can result in increased levels of anxiety, depression, and stress.
➢ Returning to face-to-face classes can also result in increased levels of anxiety, depression, and stress.

**What are the new findings?:**

➢ The prevalence of anxiety ranged from 10% to 49.4%, with higher rates recorded in female teachers, with comorbidities and working in schools.
➢ The prevalence of depression ranged from 15.9% to 28.9%, with higher rates identified in school teachers.
➢ The prevalence of stress varied from 12.6% to 50.6%, with higher rates observed among female teachers and those with chronic diseases.
➢ The only study that performed data collection during the return to face-to-face classes registered a higher prevalence of anxiety and stress than the other studies, in which the research was carried out during remote classes.

**How might this impact policy or clinical practice in the foreseeable future?:**

➢ Better training of teachers to handle the remote education model can contribute to preventing work overload and mental problems. Further, pedagogical and psychological support, especially for those who work in schools, can also prove effective.
➢ The return to face-to-face classes can increase stress and anxiety. Ensuring bio-safety protocols for safe return to face-to-face activities, can contribute to mitigating anxiety and stress about the risk of contracting the disease.
➢ There is insufficient evidence to determine a cause and effect relationship of the COVID-19 pandemic with anxiety, depression, and stress among teachers. Prospective cohort studies with control of confounding factors are necessary to infer that the pandemic has increased mental health problems in these professionals.

## INTRODUCTION

During the COVID-19 pandemic, countries have implemented measures of social distancing as a strategy to reduce the speed of spread of the contagion and to organize health services for the care of infected patients.[1] In this context, face-to-face classes have been suspended in most countries, with remote classes taking their place.[2,3] Data from the United Nations Educational, Scientific and Cultural Organization,[4] reveal that more than 190 countries have closed schools nationwide during the COVID-19 pandemic.

Although the use of online learning resources is already common, especially in the university community, the new reality of 100% remote classes may have had an impact on teachers’ mental health.[5] This is because the change took place in an abruptly short period of time, without adequate training for the use of digital resources, as well as, in most cases, without the provision of adequate equipment for remote classes.[6] In distance education (DE) courses, for example, classes are usually prepared by a team of professionals that includes content teachers and specialists in educational technologies, who are responsible for producing materials in a format accessible to various equipment, as well as being visually attractive.[7]

In this context, teachers highlight several challenges related to remote classes, but mainly related to didactic organization, in order to improve their online educational experience.[8,9] In addition, another potential risk factor for anxiety, stress and depression stems from the unequal access to computer equipment among students, which can compromise their participation in school activities. Worldwide, half of the students do not have access to a computer, and approximately 40% do not have access to the internet.[10] Thus, depending on the context, it may be necessary to adopt several strategies to assist students who do not have regular access to the internet, by providing them with printed materials and engaging them in activities. This could in turn overload teachers and increase the risk of mental illness.[11]

In the context of major changes in the professional practice of teachers, monitoring of mental health is important. Studies carried out during the COVID-19 pandemic have recorded different levels of anxiety and depression among teachers.[12,13] However, to the best of our knowledge, there is still no systematic review on this topic in the literature. The objective of this review is to identify the prevalence of anxiety, depression, and stress among teachers at schools and universities during the COVID-19 pandemic.

## METHODS

A systematic review of observational studies elaborated according to the recommendations of the *Preferred Reporting Items for Systematic reviews and Meta-Analyzes* (PRISMA) was conducted.[14] The protocol was registered at PROSPERO under number CRD42021240543.

### Research question

What is the prevalence of anxiety, depression, and stress among teachers during the COVID-19 pandemic?

### Inclusion criteria

Original studies, published between 2020 and 2021 in any language, which met the following criteria, according to the acronym PICoS, were considered eligible:

Population (P): Nursery, pre-school, elementary, high school, or higher education teachers;

Interest (I): Anxiety, depression, and stress;

Context (Co): COVID-19 pandemic;

Study type (S): Observational studies (cross-sectional, case-control, and cohort);

### Exclusion criteria

Report and case series, randomized clinical trials, literature reviews, books, and conference abstracts were excluded.

### Database search

The virtual search was carried out in the databases Pubmed, Embase, PsycINFO, Web of Science, LILACS and SciELO, using the descriptors teacher, faculty, professor, anxiety, depression, stress, insomnia, COVID-19, and their synonyms. Chart 1 shows the descriptors used in the search strategy for each database.

**Chart 1.** Databases and search strategy used to identify the studies.

| Data base | Search strategy | Applied filters |
| --- | --- | --- |
| Pubmed, Embase, Web of Science, PsycINFO | (prevalence OR "prevalence studies" OR "prevalence study" OR "cross-sectional study" OR "cross-sectional studies" OR "cross-sectional survey" OR "cross sectional analysis" OR "cross sectional analyzes" OR "disease frequency surveys" OR cohort OR "cohort study" OR "cohort Analysis" OR "cohort analysis" OR "follow-up studies" OR "longitudinal studies" OR "prospective studies" OR "retrospective studies") AND (anxiety OR nervousness OR "anxiety disorders" OR "social anxiety" OR "anxiety neuroses" OR "anxiety disorder" OR "neurotic anxiety state" OR depression OR depressions OR "depressive symptoms" OR "depressive symptom" OR "emotional depression" OR "emotional depressions" OR stress OR "stress, psychological" OR "psychological stresses" OR "life stress" OR "life stresses" OR "psychologic stress" OR insomnia OR "sleep initiation and maintenance disorders" OR "sleep initiation dysfunction" OR sleeplessness OR "insomnia disorder" OR "insomnia disorders" OR "psychophysiological insomnia" OR "sleep disturbance" OR "sleep disturbances" OR "sleep problems" OR "sleep disorders" OR "mental health") AND ("COVID-19" OR "SARS-CoV-2" OR "coronavirus disease 2019" OR "coronavirus infection" OR "2019 novel coronavirus infection" OR "2019-nCoV disease" OR "COVID19" OR "severe acute respiratory syndrome coronavirus 2" OR "coronavirus disease-19" OR "2019 novel coronavirus disease") AND (teacher OR teachers OR "school teacher" OR "school teachers" OR teacher OR faculty) NOT ("systematic review" OR "narrative review" OR "meta-analysis" OR "case-control" | No filter was applied to any of the databases. |
|  | OR "Randomized controlled trial" OR "clinical trial") |  |
| LILACS<br>and<br>SciELO | (anxiety OR depression OR stress OR insomnia) AND ("COVID-19" OR "COVID19"<br>OR "SARS-CoV-2" OR coronavirus) AND (teacher) | No filter was<br>applied to<br>any of the<br>databases. |

### Screening and selecting records

The screening of the records identified in the databases was carried out independently by two researchers (DS and RC) by reading the title and abstract. Then, the full text was independently analyzed by two researchers (DS and RC) to verify whether the studies met the inclusion criteria.

### Data extraction

Data extraction was performed by two authors (DS and RC) in an electronic spreadsheet containing the following fields: authorship, year of publication, country of study, sample characteristics, including number, age and gender, criteria for assessing anxiety, depression and stress, prevalence anxiety, depression and stress, and the prevalence by sex and place of work (schools or university).

### Evaluation of the methodological quality of the studies

The Newcastle-Ottawa Scale (NOS) modified for cross-sectional studies was used to assess the methodological quality of the studies included in the review.[15] Although this scale has seven items distributed in the domains of selection (4 items), comparison (1 item) and outcome (2 items), in this systematic review, only the following were analyzed: sample representativeness, sample size, response rate, evaluation outcome and statistical analysis, since the items exposure and comparability were not applicable to the type of study included. The total score for each study can vary from 0 to 6, because for each item a star can be assigned and for the outcome item up to two stars can be assigned. Thus, the classification of the methodological quality of the studies was carried out considering the total number of points received: ≥ 4 – good quality and <4 – low quality.

### Data analysis and results synthesis

Considering the small number of studies and the heterogeneity in the evaluation criteria and/or cut-off points used, a narrative synthesis was carried out on the prevalence of anxiety, depression, and stress among teachers during the COVID-19 pandemic. The recommendations of the synthesis without meta-analysis (SWiM) in systematic reviews were also considered.[16] With this, the number of studies that found a higher prevalence of anxiety in women was counted and compared with the number of studies that identified a higher prevalence in men. For the place of professional practice (school vs university), the same strategy was used for the synthesis of the data.

## RESULTS

The search in the databases retrieved 1,370 records and 2 were identified through manual search and analysis of article references, totaling 1,372 records. With the initial analysis of eligibility by reading the title and abstract, 42 articles were selected, of which 20 were duplicates, resulting in 22 articles for reading text-complete. Six studies, with 91,508 teachers, met the eligibility criteria and were included in the review.[12,13, 17–20] Figure 1 shows the flowchart of the study selection process for inclusion in the systematic review.

**Figure 1.**
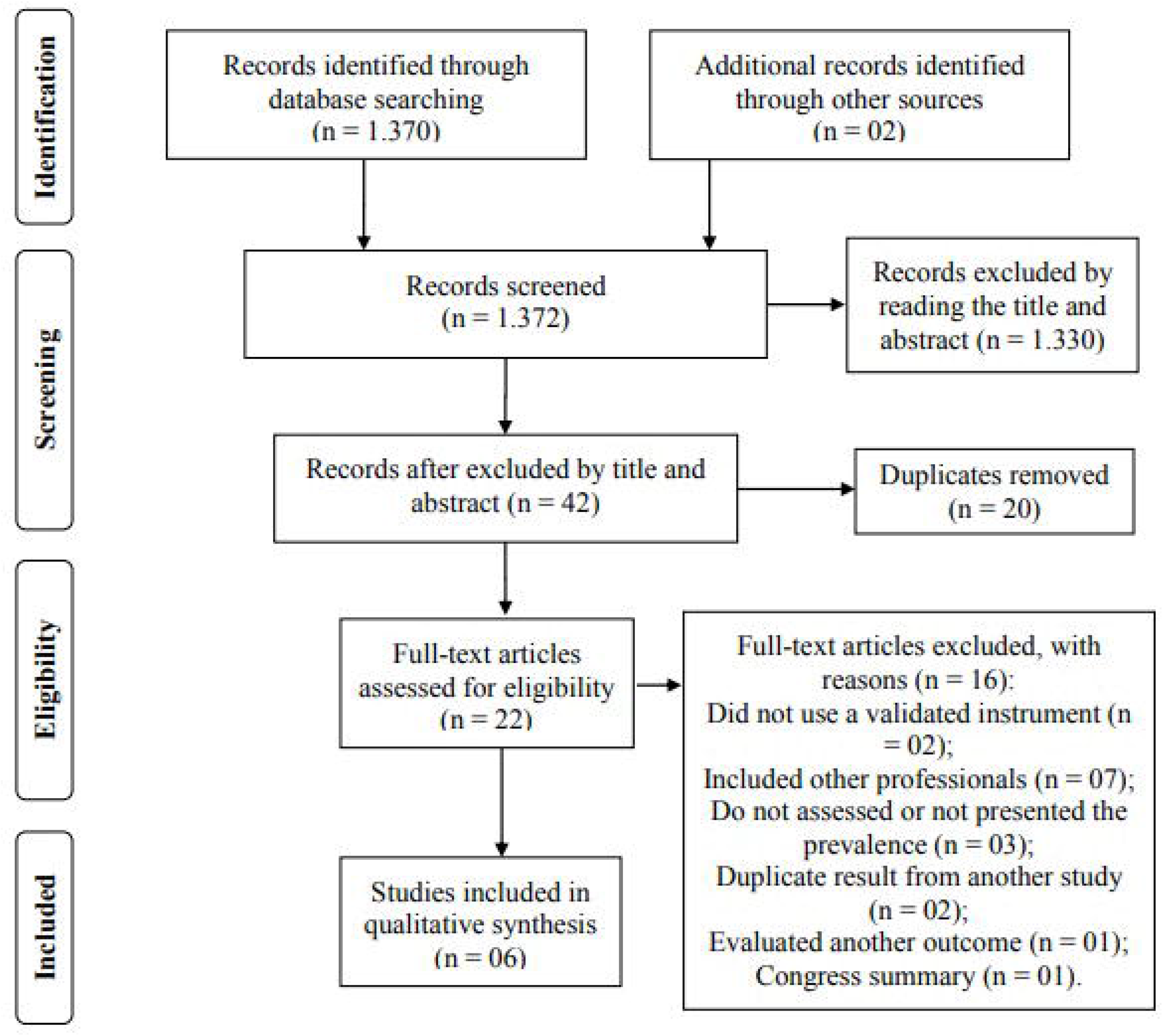
PRISMA flow chart for selection of studies.

### Characteristics of the studies

All studies presented a cross-sectional design.[12,13,17–20] Regarding the country of performance, two were in China,[17,18] one in Brazil,[12] one in the United States of America,[19] one in India[20] and one in Spain.[13] In five studies, more women than men were included.[12,13,17–19] In the studies, the ages ranged from 24[12,20] to 60 years.[20] However, there were studies that reported only the average age, which ranged from 36[17] to 42 years.[13]

Three studies included only school teachers,[12,18,20] two included school and university teachers,[13,17] and one only university teachers.[19] Only one study referred to the academic training of teachers. The majority (64.95%) held a bachelor’s degree, 23.10% had a high school degree, 6.65% held a master’s degree and the rest had other levels of education.[17] Regarding the form of professional performance, in four studies[12,17,18,20] all dealt with remote activities. One study[19] reported that 60.6% of teachers were engaged in remote activities and one study included teachers who returned one to two weeks before (this study date) for face-to-face classes.[13] Table 1 presents the characteristics of the studies included in the review.

**Table 1.**
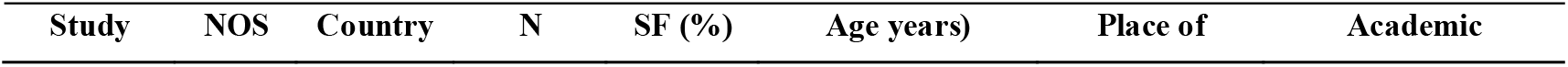
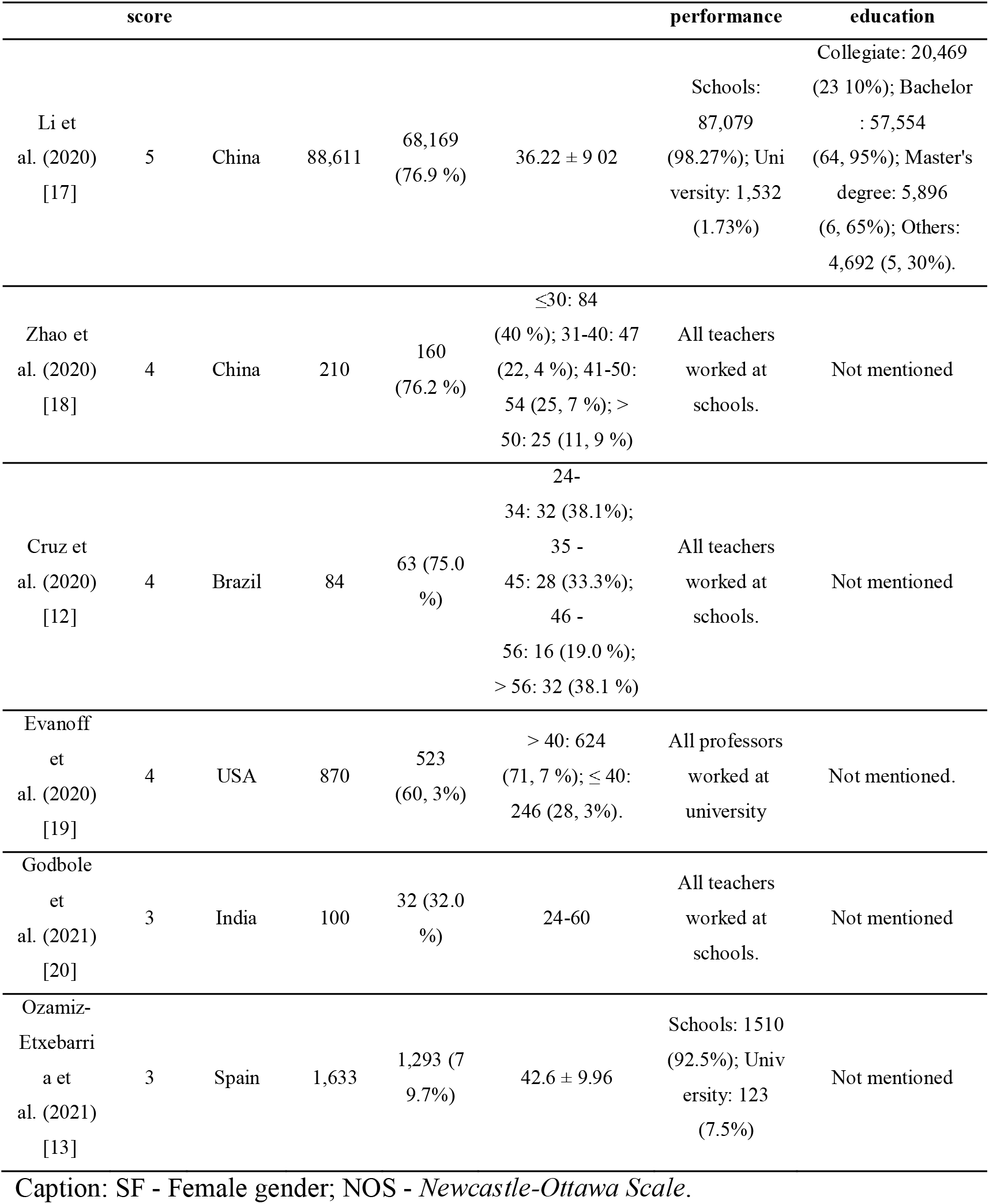
Characteristics of the studies included in the review (n = 6)

Regarding exposure to COVID-19, the study by Cruz et al. (2020), with Brazilian teachers, recorded that 15.5% of the participants reported contact with a COVID-19 positive person.[12] Evanoff et al. (2020) found that 16.3% of teachers reported having experienced some exposure to COVID-19. [19] No study reported diagnosis of teachers with COVID-19. Only the study by Ozamiz-Etxebarria et al. (2021) presented data on chronic diseases in the participants, recording that 16.7% had at least one.[13]

### Criteria for assessing anxiety, depression, and stress

Three studies included in the review used the Depression, Anxiety and Stress Scale-21 Items (DASS-21) for the diagnosis of anxiety, depression, and stress.[12,13,19] In two, the standard cutoff points of the instrument were used: 5 for mild, 10 for moderate and 15 for severe,[13,19] and one study did not mention the cutoff point.[12]

Three studies used criteria other than DASS-21 to assess anxiety. The Generalized Anxiety Disorder tool (GAD-7) was used in the study by Li et al. (2020),[17] with a cut-off point of 10, referring to moderate or severe anxiety. Zhao et al. (2020) used the Self-Rating Anxiety Scale (SAS), using 50 as the cutoff point.[18] Godbole et al. (2021) used the Hamilton Anxiety Rating Scale, with a cutoff point of 18-24 for moderate anxiety and 25-30 for severe anxiety.[20]

### General prevalence of anxiety, depression, and stress

The prevalence of anxiety among teachers during the COVID-19 pandemic ranged from 10%[19,20] to 49.4%.[13] In the two studies where the assessment was on the bases of sex,[13,17] a higher prevalence of anxiety was recorded in females. Ozamiz-Etxebarria et al. (2021) reported a higher prevalence of anxiety in teachers with chronic diseases (61.9% vs 47.0%).[13] Two studies found that teachers with older chronological age had a lower prevalence of anxiety[13,19] and Li et al. (2020) found no difference in the average age between teachers with anxiety and without anxiety.[17] In the only study that showed the prevalence of anxiety by degree of academic education, a similar prevalence was found, with 14.7% among high school teachers, 12.7% among those with bachelor’s degrees and 13.4% for those with master’s degrees.[17]

Regarding the prevalence of anxiety by place of professional activity, among teachers who work in schools it ranged from 10%[20] to 21.7%,[12] while for teachers employed at a university it ranged from 10%[19] to 12.9%.[17] Reviewing the data by continent, in studies carried out in Asia the prevalence of anxiety ranged from 10%[20] to 17.2%,[18] in the Americas from 10%[19] to 21.7% and in Europe 49.4%.[13]

The prevalence of depression ranged from 15.9%[19] to 28.9%[12] in studies conducted in the Americas and the study conducted in Europe identified a prevalence of 32.2%.[13] In the study that showed prevalence by sex, no significant difference was identified.[13] The prevalence of depression was similar between age groups in one study.[13] Another study found that teachers aged over 40 years had a lower prevalence rate for depression.[19] Ozamiz-Etxebarria et al. (2021) reported a higher prevalence of depression in teachers with chronic diseases (41.0% vs 30.4%).[13] The study carried out with university professors[19] identified a considerably lower prevalence than that carried out with school teachers.[12]

The prevalence of stress ranged from 12.6%[19] to 12.7%[12] in studies in the American countries. The European study registered a prevalence of 50.6%.[13] Ozamiz-Etxebarria et al. (2021) identified a prevalence of stress in females of 52.1% and 43.9% in males.[13] In the study by Ozamiz-Etxebarria et al. (2021)[13] teachers aged over 46 years had a lower prevalence of anxiety. Similar results were presented by Evanoff et al. (2020).[19] A higher prevalence of stress was found in teachers with chronic diseases (71.4% vs 64.1%).[13] No study reported the prevalence of anxiety, depression, and stress among teachers, in the context of exposure to COVID-19. Table 2 shows the prevalence of anxiety, depression, and stress reported in each study.

**Table 2.**
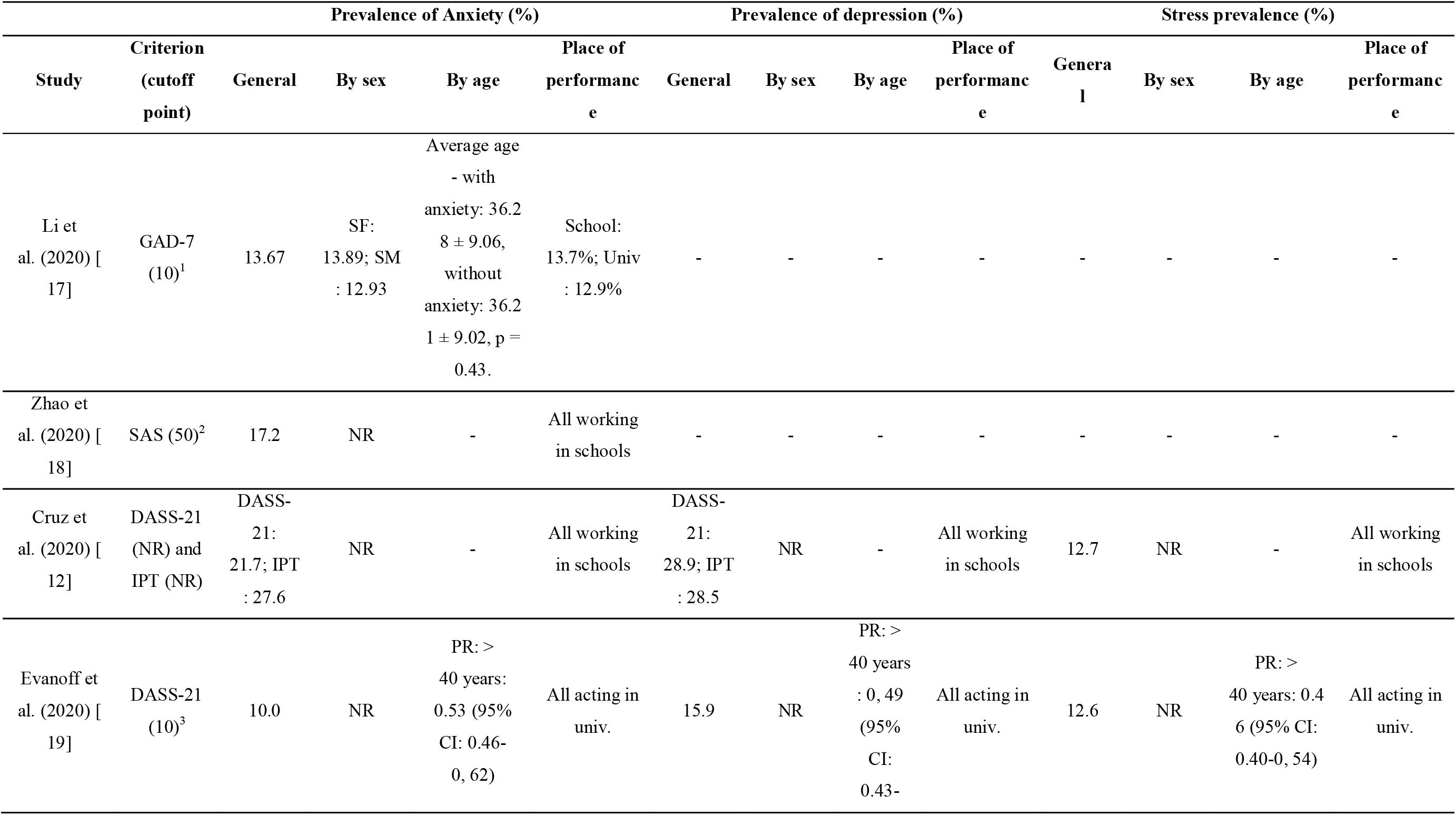
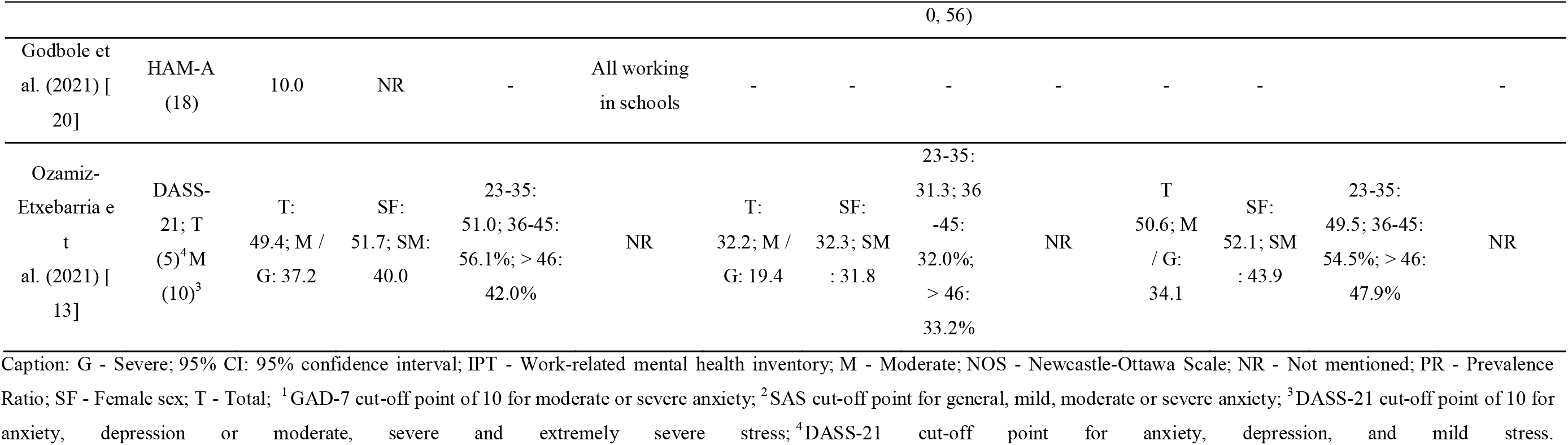
Prevalence of anxiety, depression, and stress in studies with teachers during the COVID-19 pandemic (n = 6)

## DISCUSSION

To the best of our knowledge, this is the first systematic review to assess the prevalence of anxiety, depression, and stress in school and university teachers during the COVID-19 pandemic. The prevalence of anxiety ranged from 10% to 49.4% and was considerably higher in the study conducted in Europe. The prevalence of depression ranged from 15.9% to 28.9%, being considerably higher in the study with teachers who worked in schools. For stress, a considerably higher prevalence was found in Europe (50.6%) than in studies conducted in the Americas (12.7%).

The study conducted in Spain (Europe)[13] recorded a considerably higher prevalence of anxiety and stress compared to the other studies and was the only one where, during data collection, teachers had returned to face-to-face classes, after a period of remote classes. The higher prevalence of anxiety and stress can be explained, in part, by the uncertainty of the impact of face-to-face classes on the risk of contagion, due to the greater need for commuting, as well as by the possibility of greater contact with other professionals from schools or universities, as well as with students.[21] In addition, the return to face-to-face classes with strict bio-safety protocols and the teachers’ “enhanced responsibility to monitor the students” may be related to a higher prevalence of anxiety and stress.[2,22]

Another factor that could contribute to increased levels of anxiety and stress when returning to in-person classes is the health status of teachers; there was a higher prevalence of anxiety in teachers with chronic diseases.[13] Several studies have recorded a high prevalence of obesity, hypertension, diabetes, respiratory disorders and other chronic non-communicable diseases in teachers.[23–25] Considering that obesity, diabetes and hypertension are associated with higher mortality due to COVID-19, when resuming classes, teachers with chronic diseases may be more afraid that, if infected, they could see more harmful effects. This could justify the greater anxiety among this group.[26]

In this context, it is important that prior to the reopening of schools, education professionals receive training in the measures to be taken to minimize the risk of infection due to COVID-19. This, in addition to helping reduce the incidence of cases among the school or university community, can contribute to reducing the degree of anxiety and stress when returning to face-to-face classes. Li et al. (2021), in a study with 67,357 teachers in China, found that the lack of knowledge about the proper type of mask and the correct way to use it, as well as the non-adherence to the use of a mask, were factors associated with a higher risk of anxiety.[27] In addition, it is important to provide good quality personal protective equipment (PPE) and in sufficient quantity. A study conducted with 2,665 teachers in Denmark found that the shortage of PPE, as well as greater contact with parents of students and other education professionals, were factors associated with more frequent changes in emotional state.[28]

Regarding the prevalence of anxiety according to the place of professional activity, a higher prevalence of anxiety was found among school teachers[12,18] as shown in two studies, than among university professors. One possible explanation is that school teachers may have less experience in remote education than those at university.[29] In this sense, a study carried out during the COVID-19 pandemic with 260 school teachers in the United States of America, a developed country, registered that 52% referred to the challenge of scarcity or little knowledge about strategies for remote/online education and 44% were unaware of the communication tools required for remote/online classes.[30]

Another hypothesis for this difference in relation to the place of work may be that university professors engage with young adults who may find it easier to adapt to remote education than children and adolescents, who represent the majority of the students of school teachers.[31,32] In addition, studies conducted during the pandemic have found that parents have devoted considerable time (more than 1 hour a day) to assisting their children in remote classes.[32,33] With regard to school teachers, in addition to online teaching strategies, there is a need for strategies to facilitate communication with the children’s parents. This is an additional task and could be one more factor that adds to anxiety and stress.[30]

Regarding depression, teachers working at schools also had a higher prevalence than those working in universities. As university professors are generally more accustomed to remote education, this may have favored the lower prevalence of depression in this group.[29] In addition, as the study with school teachers was carried out in Brazil and with university teachers in the United States of America, a possible explanation for this result is the lower remuneration among Brazilians, which forces teachers to do more than a single job. This has been associated with a higher risk of depression.[34,35] In this sense, Patel et al. (2018), in a meta-analysis of 12 studies, found that there is a greater risk of depression in countries with higher income inequality than in countries with lower income inequality.[36]

Another probable explanation for depression being less prevalent among university professors is the fact that these professionals generally have a higher degree of academic training. In Brazil, for example, 4.6% of Basic Education teachers (schools) had a master’s degree or doctorate in 2017,[37] while 64.3% of university professors had completed a doctorate.[38] Data from the United States Department of Education revealed that in 2018, just over 50% of primary and secondary school teachers completed a master’s degree or doctorate,[39] while for university professors this percentage was 68%.[40] In this context, studies have found that a higher degree of academic training is associated with a lower risk of depression, possibly mediated by greater financial stability, which may represent better access to health services, as well as may influence the adoption of behaviors beneficial to mental health.[41.42]

Regarding sex, there was a considerably higher prevalence of anxiety and stress among females than males.[13,17] The first explanation for this may simply be the greater female participation in the studies included in the review. However, studies have found that women are at higher risk for anxiety and stress, which may be related to high levels of estrogen and greater sensitivity to increased catecholamine in the consolidation of emotional memory.[43,44]

In addition, the greater participation in housework by women, as well as the greater investment of time to help their children with schoolwork, are factors that can contribute to a higher prevalence of anxiety and stress in women.[45] These factors can also contribute to gender inequality in the academic production of female teachers, including during the COVID-19 pandemic, as many university teachers are also researchers.[46] In this sense, Gabster et al. (2020), in an analysis of 1179 articles on COVID-19, found that in 28% and 22% of the studies, the first author and the last author were women, respectively. In comparison with articles published in the same journals in 2019, a reduction of 23% and 16% of women as first author and last author (respectively) was seen.[46]

There was a considerably lower prevalence of anxiety and stress among older teachers compared to younger teachers in the two studies.[13,19] Among the general population, a higher prevalence of anxiety was seen among younger people during the COVID-19 pandemic.[47] Older people may have a lower risk of anxiety and stress due to possible greater resilience gained from exposure to various stressful situations over time, which may favor better emotional control.[47,48]

This study, to the best of our knowledge, is the first systematic review to assess the prevalence of anxiety, depression, and stress among teachers during the COVID-19 pandemic. In addition, the search for studies was carried out on six databases, in order to seek comprehensive coverage of the literature on the topic and the narrative synthesis following the pattern recommended by SWiM.[16] However, this review has some possible limitations, such as the limited number of studies and the high heterogeneity among them, which limited us to carry out the meta-analysis. Furthermore, considering the cross-sectional nature of the included studies, it is not possible to infer causality in factors related to anxiety, depression, and stress. Regarding the prevalence of anxiety, the different diagnostic criteria and cutoff points used in the studies may have influenced the different prevalence levels. Finally, it was not possible to separate the analysis of prevalence among teachers from public and private schools where differences in structure, remuneration and teaching resources could influence the prevalence of anxiety, depression, and stress. The results cannot be extrapolated to countries with different cultures, economies and educational systems that were not included in the studies considered in this review.

In addition, the review has practical implications, as it indicates the need for better training of teachers to work in the remote education model, with pedagogical and psychological support that prevents work overload and mental problems. Likewise, the return of face-to-face classes can increase the prevalence of stress and anxiety, indicating that these professionals are involved in biosafety protocols for safe return to face-to-face activities. This can contribute to teachers being less anxious and stressed about the risk of contracting the COVID-19 virus.

## CONCLUSION

The prevalence of anxiety among teachers was high during the COVID-19 pandemic, varying from 10% to 49.4%, with the highest prevalence among participants in the study carried out in Spain (where face-to-face classes were witnessing a return). The rampancy of stress was also higher in the participants of the European study compared to those in the studies carried out in the Americas. Depression was more prevalent among teachers who worked in schools.

The results show a high prevalence of anxiety, depression, and stress among teachers and alerts us to the need for greater care of mental health issues. However, due to discrepant data in the different studies included in the review, studies with more rigorous methodology and standardization of diagnostic instruments are necessary to know the real impact of the pandemic on the mental health of these professionals.

## Data Availability

Data available on reasonable request.

